## Supplementary Information for "Convergent trends and spatiotemporal patterns of arboviruses in Mexico and Central America"

**"Trends and spatiotemporal patterns of arboviral spread in Mexico and Central America": Supplementary Information**

**Supplementary Text 1**

***Retrospective on the arbovirus epidemics in the Americas and epidemiological surveillance in Mexico***

*Chikungunya virus (CHIKV)*

The first locally acquired infections were reported in the Caribbean islands in 2013 [74,75], with subsequent spread into Central and South America during 2014[65,70,71]. The outbreaks in the Caribbean islands affected various territories over 2014[94], and reached high incidences in Guadeloupe (295.8 cases per 1000 inhabitants in November 2014), the French Guiana (112.41 cases per 1000 inhabitants in July 2014) and the Dominican Republic (over 142 000 new cases reported in July 2014). CHIKV reached South America and the USA by June 2014, followed by Mexico and various other countries in Central America by the end of that year [95]. Most of these outbreaks have been attributed to a single introduction of the Asian genotype into the Eastern Caribbean [72], except for Brazil, where a secondary and independent introduction event of the Eastern/Central/South African (ECSA) virus genotype was identified[63]. The ECSA genotype has since then spread across Brazil [44,73], and was recently introduced into Paraguay [96]. It is believed that the spread of CHIKV in the region was at least partially driven by international travel [69,97,98], whilst a successful virus establishment was likely aided by (*i*) the lack of prior immunity to the virus in the Americas, (*ii*) the presence of both *Ae. aegypti* and *Ae. albopictus* in the same geographical region, and (*iii*) and elevated volumes of travel across countries with high virus transmission [28]. CHIKV was detected in Mexico in 2014, with the highest number of cases reported during 2015, particularly within the southern and southwestern states of Guerrero, Michoacán and Yucatán [32]. From 2016 onwards, limited numbers of cases have been reported, although CHIKV incidence in southwestern states (such as Guerrero) is thought to be underestimated, represented by undiagnosed or non-hospitalized cases that do not make it to the SINAVE official case report list [99].

*Dengue virus (DENV)*

DENV is classified into four immunologically and genetically distinct serotypes: DENV-1, DENV-2, DENV-3 and DENV-4. Before the 1960s, DENV-2 was the only serotype detected in the Americas. Nonetheless, the other three serotypes were detected throughout the 1960s to the 1990s, including the characterization of multiple introductions of genetically distinct DENV-2 lineages [29,100]. Once established within a geographical region, DENV epidemics follow seasonal patterns (as observed in countries in the Americas) [10,48,101–103], with particularl prominence in regions where the virus has become endemic (defined as where at least one virus serotype constantly circulates [104]) or hyperendemic (defined as where multiple virus serotypes constantly co-circulating). In these scenario, seasonal cycles are characterised by yearly periods where outbreaks are more frequent, driven in part by changing climatic conditions that favour vector breeding and increased vector-host interactions[105-108]. Furthermore, the co-circulation of different DENV serotypes can lead to dominance over specific season across multiple years[78,79,109]. The processes driving t long-term dominance patterns are complex and involve climate, vector ecology, demographics and immunity of the host population factors[11,110,111]. This can lead to seasonal patterns where large outbreaks vary in size and severity (for example, with higher number of cases for dengue haemorrhagic fever, DHF) across seasons through time[10,48,101–103].

Dengue has been under surveillance in Mexico since the 1980s, and became a health concern at a national level when the incidence of DHF increased during the 1990s, associated with the spread of *Aedes* mosquito populations in Mexico [33]. Since the 2000s, the geographical distribution of Dengue has marginally expanded into other locations closer to central Mexico, such as the state of Morelos and areas of higher altitude, like the state of Guerrero [77,82]. During the early 2010s, epidemiological hotspots for Dengue were identified in and around urban areas within the southern states, where transmission occurs year-round but peaks during the rainy season [82]. The incidence of DENV serotypes has fluctuated over multiple years: during the early 2000s, DENV-1 and DENV-2 alternated as the dominant serotype in Mexico, with DENV-1 replacing the previously dominant DENV-2 between 2004 and 2007[77].

#### Figure S1. CHIKV epidemiological and genomic surveillance trends in the Americas

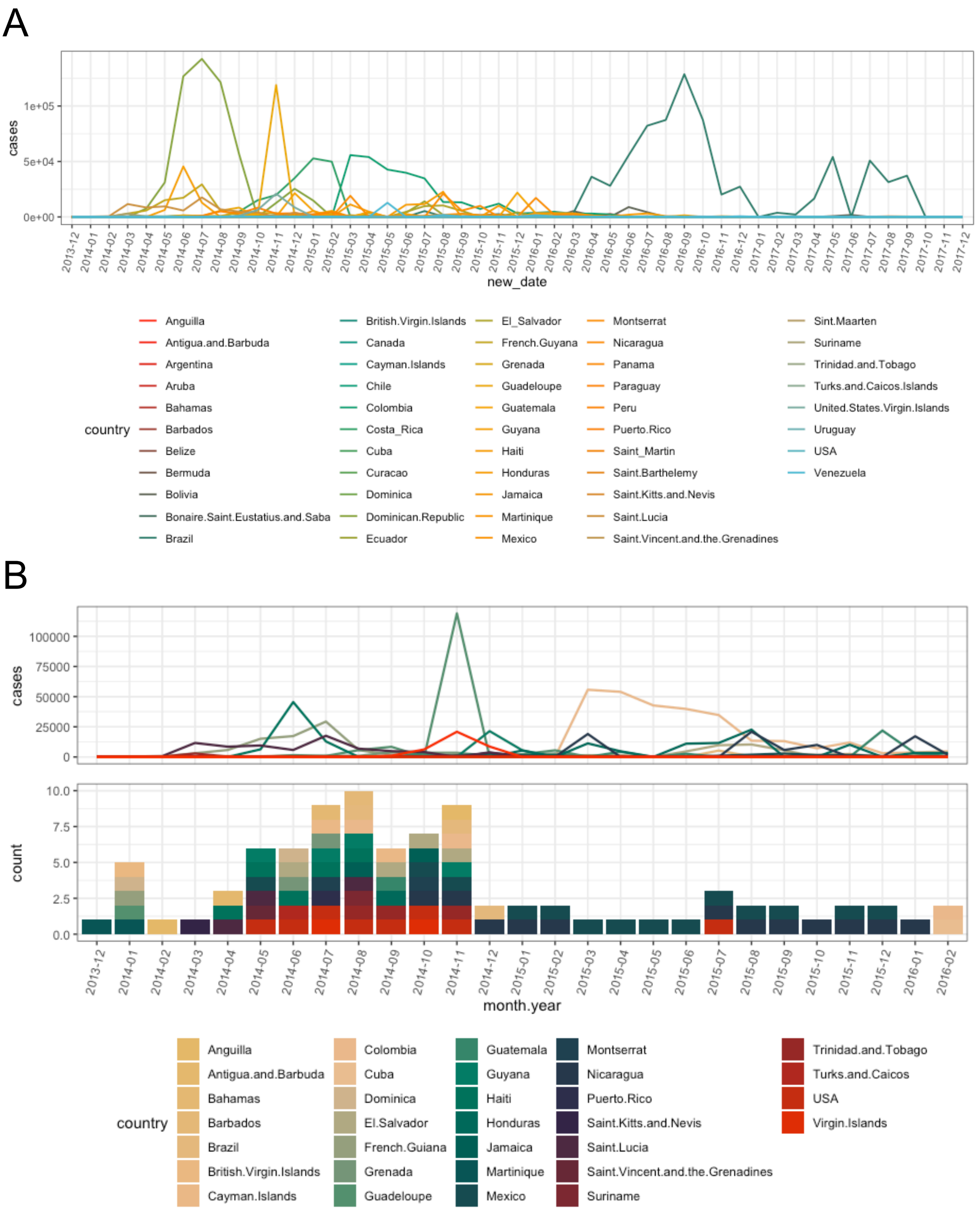


***(A)*** Monthly number of CHIKV cases reported to the Pan-American Health Organisation (PAHO) between 2013 and 2017, grouped by country. ***(B)*** Comparison between monthly number of cases (reported to PAHO, upper panel) in countries that have generated CHIKV genome sequences, and publicly available complete CHIKV genome sequences (lower panel). Mexico sequences include those generated in this study.

**Figure S2. Complete CHIKV genome sequences versus total cases reported to PAHO per country**

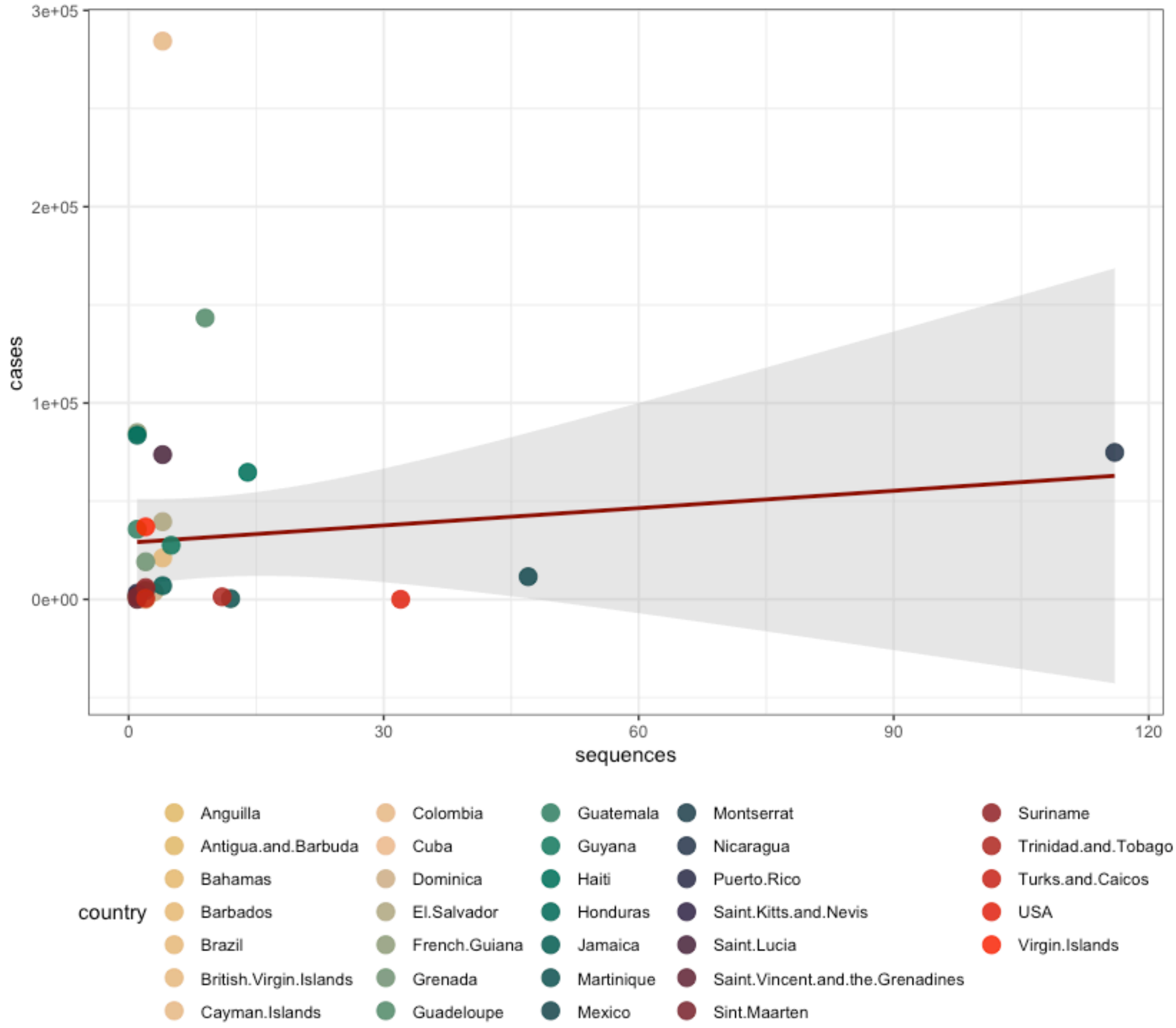


Linear regression between the numbers of CHIKV sequences and cases per country over time, with 95% confidence interval (CI) shown in grey. A Spearman's Rho = 0.26, $p$ = 0.15 denotes no correlation between the cumulative number of cases per country versus the number of viral genome sequences available per country.

**Figure S3. CHIKV epidemiological trends across Mexico between 2016 and 2018**

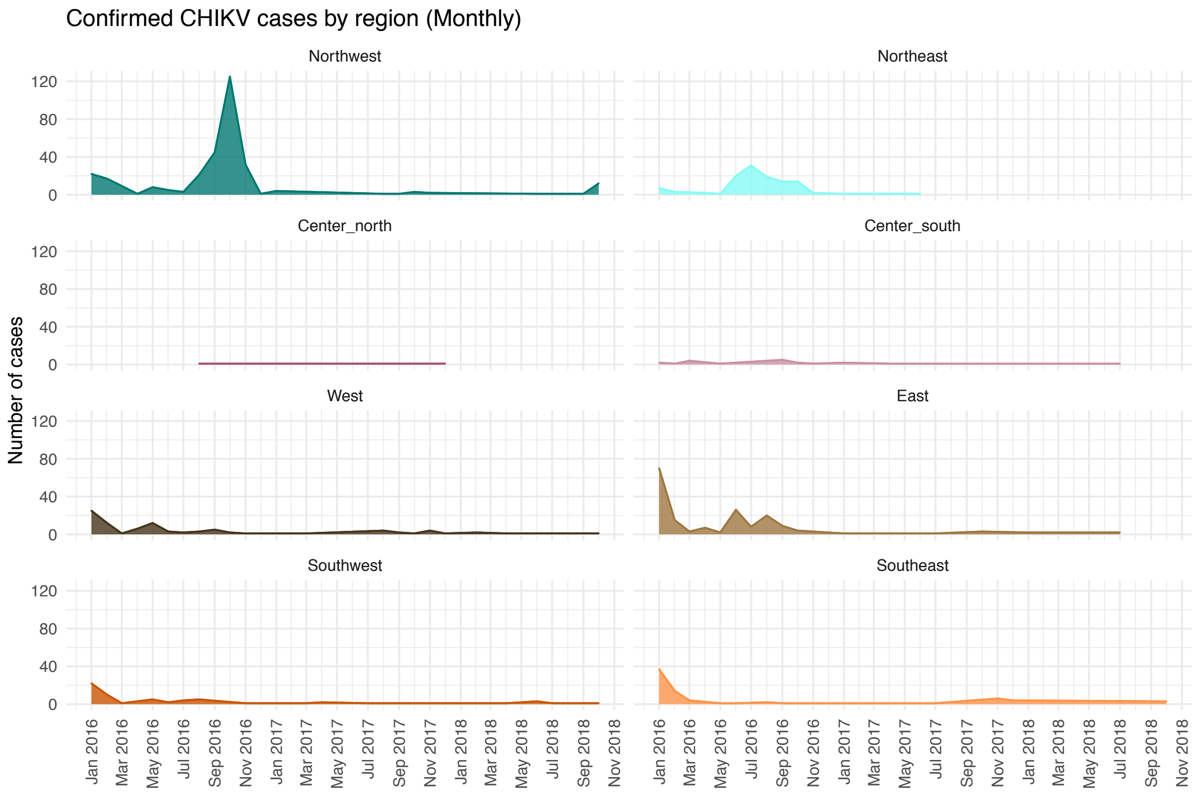


Monthly cases for CHIKV reported under the SINAVE surveillance system (InDRE/Ministry of Health Mexico).

#### Figure S4. Phylogenetic analyses of CHIKV in the Americas

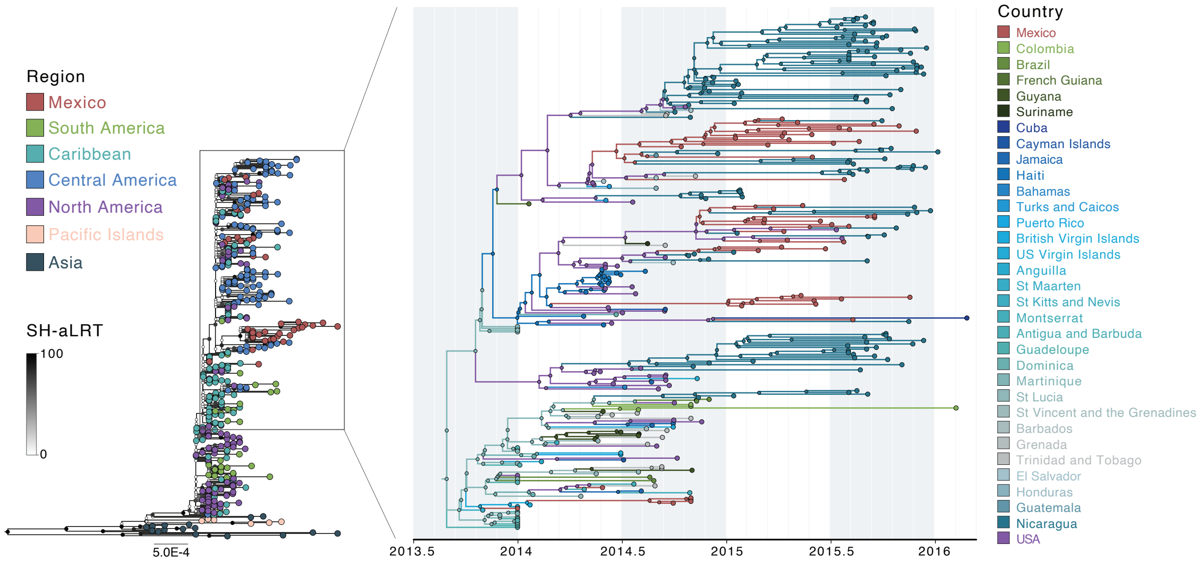


ML phylogenetic tree for CHIKV inferred from the complete genome sequences from the Americas included in our analysis, denoting an 'American' lineage (left panel). Tree tips are coloured according to the region/country of collection, whilst nodes are coloured according to branch support values (SH-aLRT). To the right, a time-calibrated MCC tree is displayed, showing a well-defined CCNA clade within the 'American' lineage. Tips and branches are coloured by the location of origin and circulation, inferred through a DTA phylogeographic analysis (see Methods section, main text).

**Figure S5. DENV epidemiological trends across Mexico between 2016 and 2018**

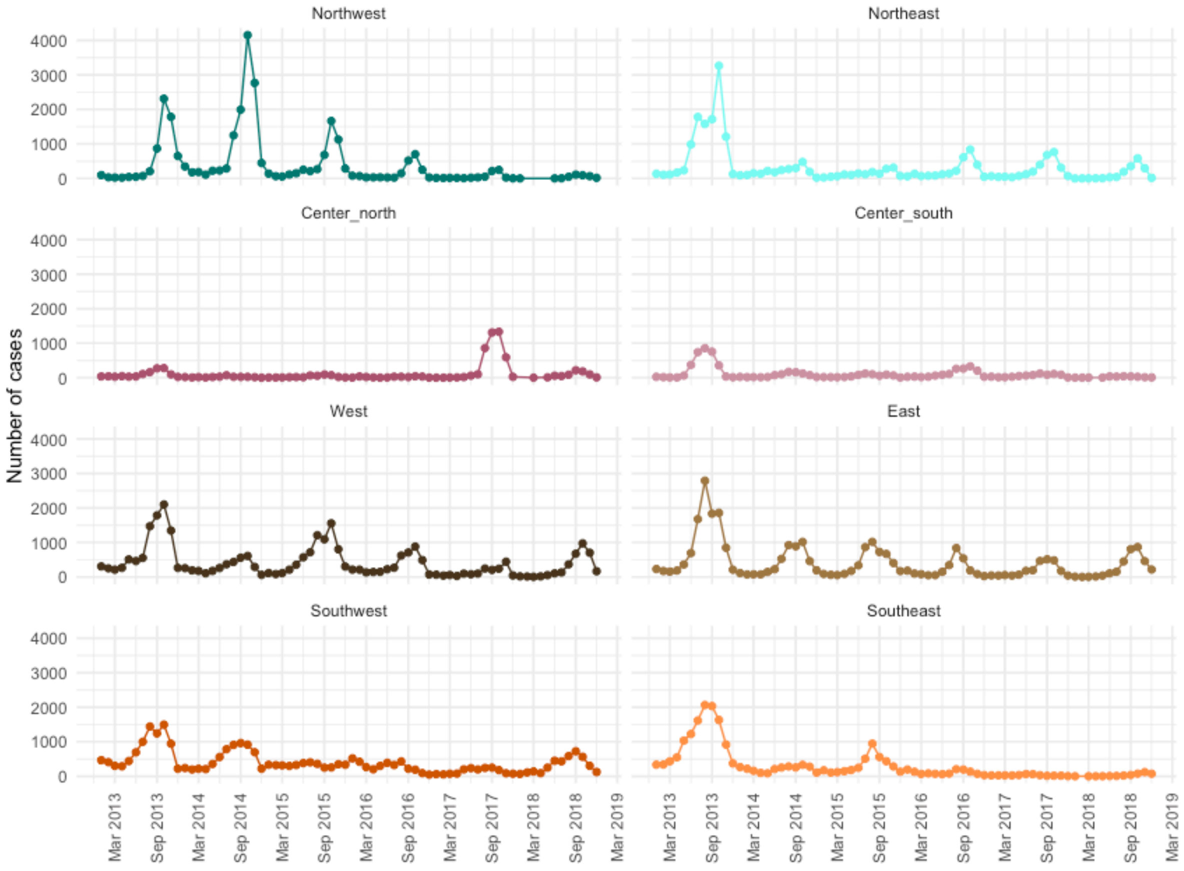


Monthly cases for DENV (aggregating both dengue fever and dengue haemorrhagic fever) reported under the SINAVE surveillance system (InDRE/Ministry of Health Mexico).

**Figure S6. Serotyping representation for DENV across regions in Mexico**

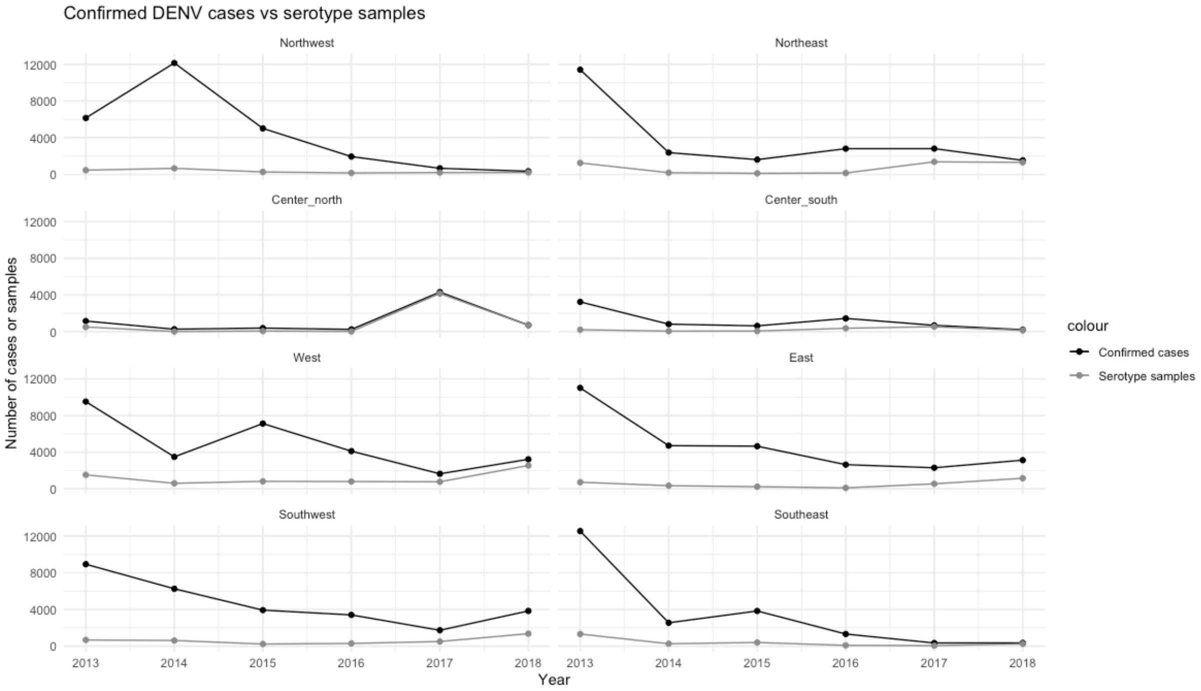


Total number of DENV cases reported by year (black line), compared to total number of DENV cases where the causal serotype has been identified and reported (grey line). Data corresponds to the period of time between 2013 and 2018, as reported under the SINAVE surveillance system (InDRE/Ministry of Health Mexico).

**Figure S7. DENV serotyping efficacy across time in different Mexico regions**

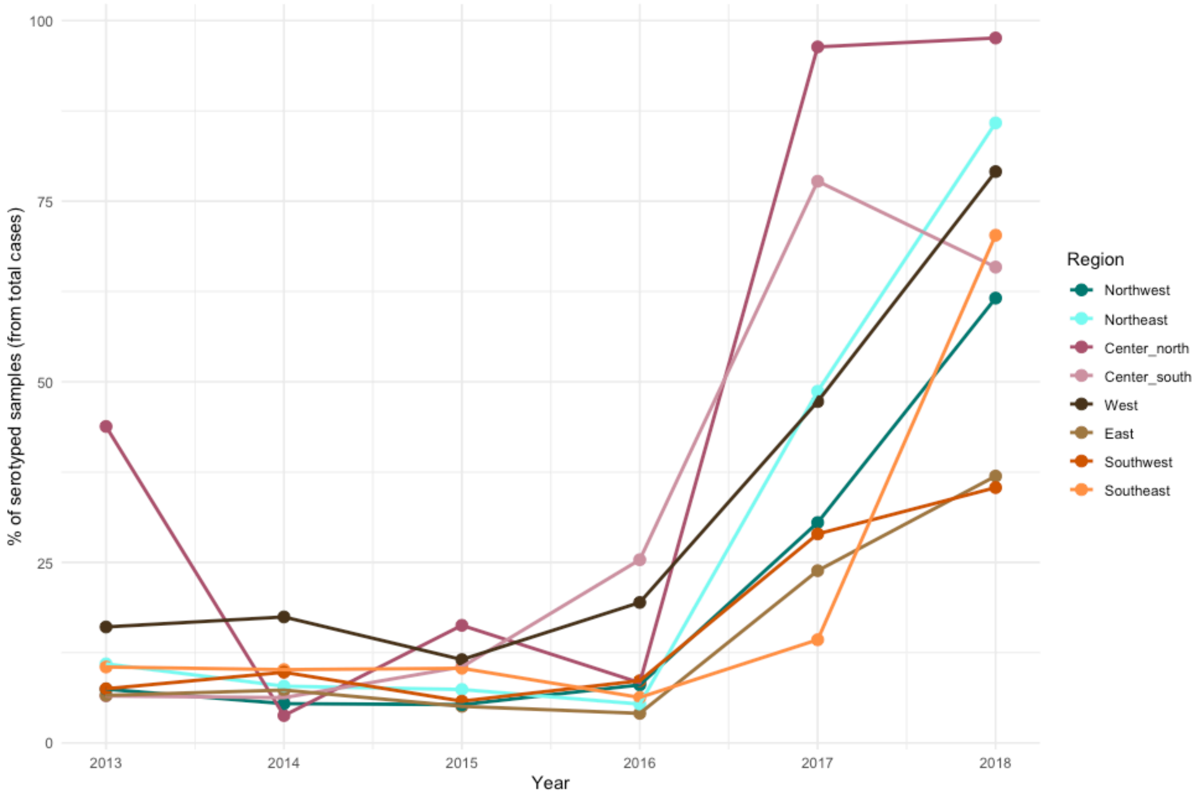


Percentage of serotyped samples per region per year between 2013 and 2018.

#### Figure S8. DENV-1 and DENV-2 case numbers across Mexico regions

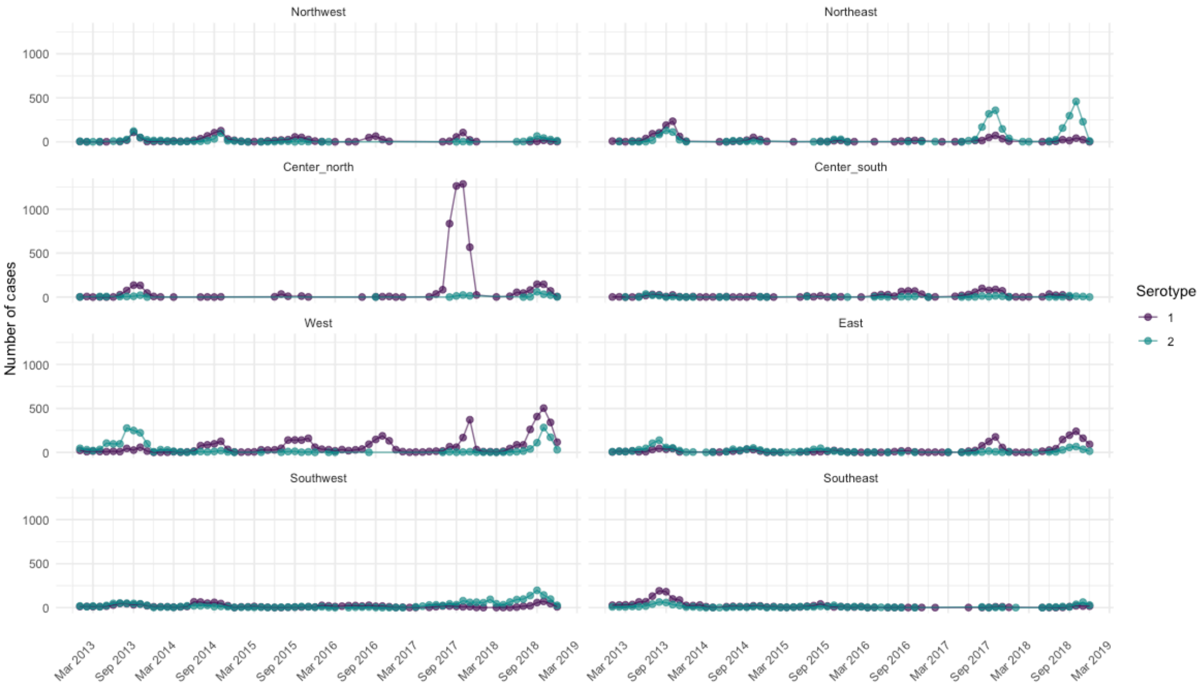


Monthly numbers of cases identified as DENV-1 (purple) or DENV-2 (teal) between 2013 and 2018 across geographic regions in the country.

**Figure S9. Phylogenetic analyses of DENV-1 and DENV-2 in the Americas**

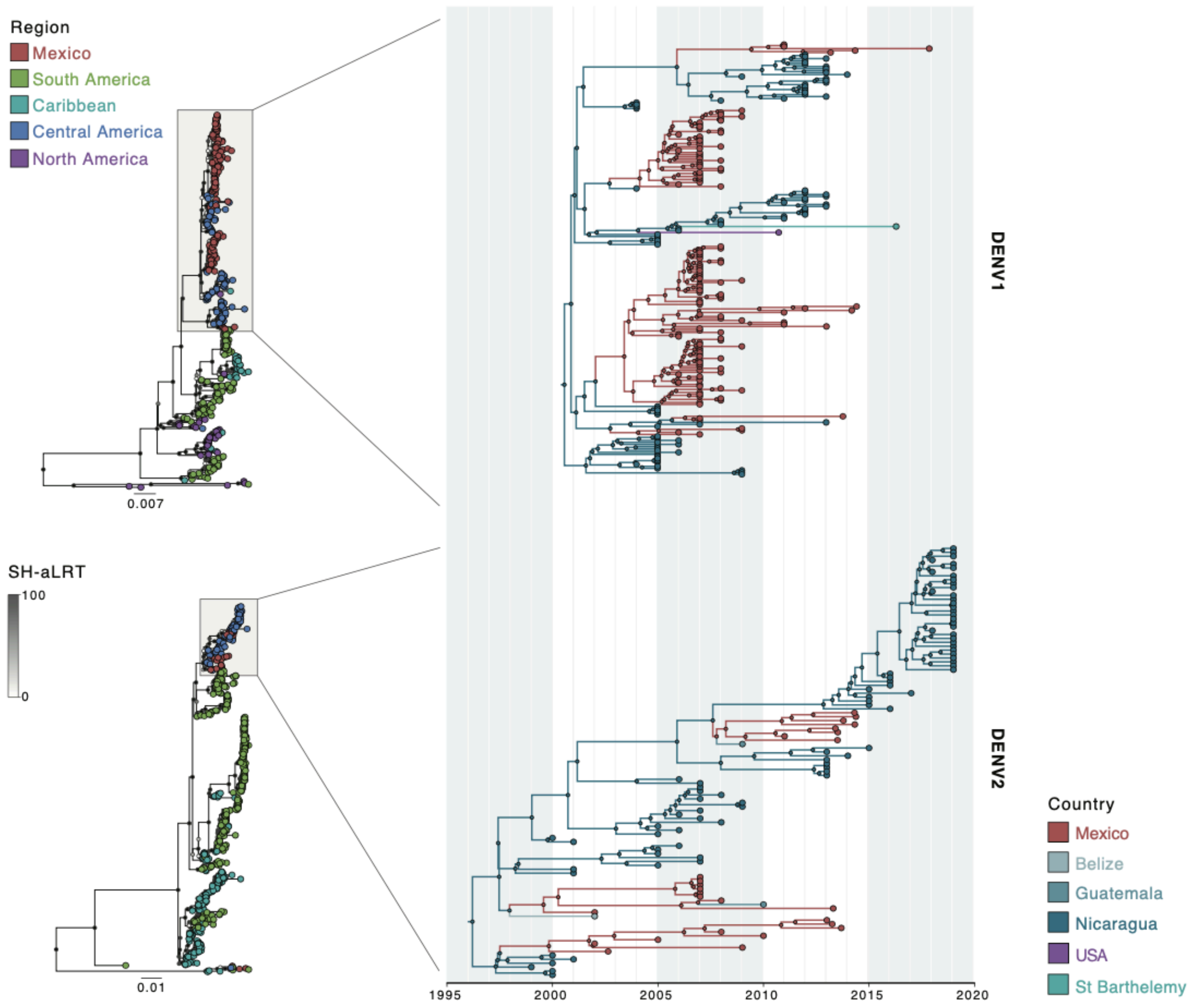


ML phylogenetic trees for DENV-1 (above) and for DENV-2 (below) inferred from the complete genome sequences from the Americas included in our analysis are shown to the left. Tree tips are coloured according to the country/region of collection, whilst nodes are coloured according to branch support values (SH-aLRT). To the right, the time-calibrated MCC trees are displayed, showing well-defined CCNA clades for each virus. Tips and branches are coloured by the location of origin and circulation, inferred through a DTA phylogeographic analysis (see Methods section, main text).

#### Figure S10. Bayesian Skyline plot of the CAM DENV-1 and DENV-2 lineages

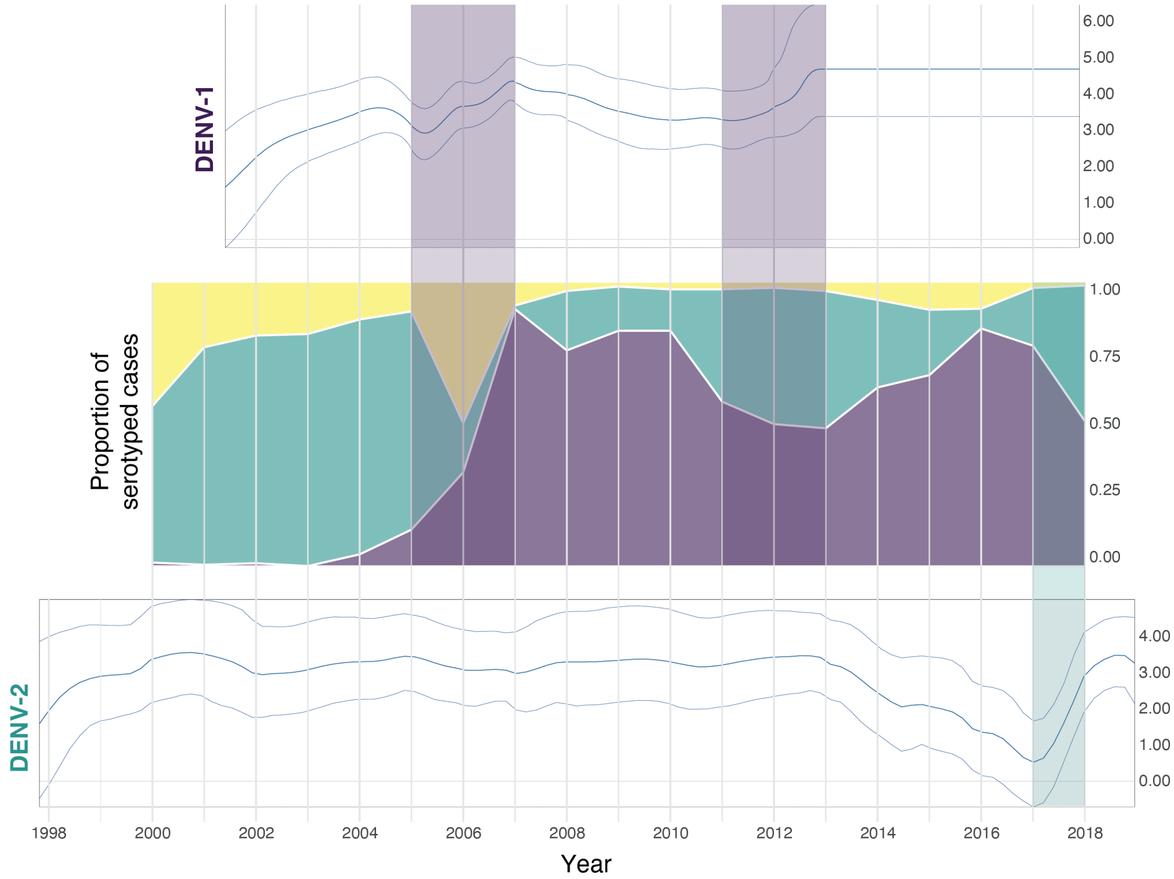


Upper and lower panels show the Bayesian Skyline plots (BSPs) obtained from the time-scaled phylogenetic analyses of the DENV-1 and DENV-2 CCNA lineages. The middle panel shows the proportion of serotyped cases for each DENV serotype in Mexico over a comparable period of time. Shading in purple (for DENV-1) and green (for DENV-2) show periods of time where an increase in the virus effective population size over time was observed (as suggested by the BSP), highlighting the proportion of virus serotypes.

### Table S1. Genome sequences generated in this study

| Virus | Genotype | InDRE ID | Host | State | Collection date |
|---|---|---|---|---|---|
| ***DENV-1*** | V | 198cc | Human | Quintana Roo | 2013-03-18 |
| | V | 112cc | Human | Veracruz | 2013-01-09 |
| | V | 321cc | Human | Chiapas | 2014-05-18 |
| | V | 640cc | Mosquito | Morelos | --- |
| | V | 1748 | Human | Jalisco | 2013-10-18 |
| | V | 3933 | Mosquito | Jalisco | 2017-11-22 |
| | V | 3761 | Mosquito | Jalisco | --- |
| ***DENV-2*** | III | 1773 | Human | Jalisco | 2013-01-02 |
| | III | 1537cc | Mosquito | Morelos | 2013-09-09 |
| | III | 649cc | Human | Quintana Roo | 2013-07-05 |
| | III | 242 | Human | Chiapas | 2014-04-25 |
| | III | 1620 | Human | Jalisco | 2013-10-15 |
| | III | 426cc | Human | Veracruz | 2013-05-30 |
| | III | 237cc | Human | Jalisco | 2013-04-04 |
| | III | 278 | Human | Colimas | 2013-04-22 |
| | III | 243 | Human | Chiapas | 2014-04-30 |
| | III | 320 | Human | Chiapas | 2014-05-19 |
| | III | 658cc | Human | Veracruz | 2013-07-10 |
| ***CHIKV*** | Asian | 5528 | Human | Baja California | 2015-12-01 |
| | Asian | 3601 | Human | Ciudad de Mexico | 2015-08-06 |
| | Asian | 169 | Human | Guerrero | 2015-01-20 |
| | Asian | 4818 | Human | Guerrero | 2015-09-18 |
| | Asian | 1118 | Human | Mexico | 2015-04-10 |
| | Asian | 2417 | Human | Michoacan | 2015-06-01 |
| | Asian | 1657 | Human | Morelos | 2015-05-07 |
| | Asian | 1479 | Human | Nuevo Leon | 2015-04-19 |
| | Asian | 2090 | Human | Queretaro | 2015-06-02 |
| | Asian | 3588 | Human | Quintana Roo | 2015-07-27 |
| | Asian | 2454 | Human | Sinaloa | 2015-06-16 |
| | Asian | 3584 | Human | San Luis Potosi | 2015-07-19 |
| | Asian | 2646 | Human | Tabasco | 2015-06-25 |
| | Asian | 741 | Human | Tampico | 2015-03-10 |
| | Asian | 4823 | Human | Veracruz | 2015-09-19 |
| | Asian | 2938 | Human | Yucatan | 2015-07-06 |
| | Asian | 3655 | Human | Zacatecas | 2015-08-05 |
| | Asian | 222 | Human | Morelos | 2015-01-29 |
| | Asian | 504 | Human | Nuevo Leon | 2015-02-27 |
| | Asian | 511 | Human | Michoacan | 2015-02-16 |
| | Asian | 698 | Human | Veracruz | 2015-03-09 |
| | Asian | 809 | Human | Jalisco | 2015-03-16 |
| | Asian | 1441 | Human | Jalisco | 2015-04-22 |
| | Asian | 2870 | Human | Oaxaca | 2015-07-09 |
| | Asian | 3364 | Human | Veracruz | 2015-07-28 |
| | Asian | 4320 | Human | Morelos | 2015-08-23 |
| | Asian | 4823 | Human | Veracruz | 2015-09-19 |
| | Asian | 1449 | Human | Chiapas | 2015-04-13 |
| | Asian | 1309 | Human | Chiapas | 2015-04-09 |
| | Asian | 948 | Human | Quintana Roo | 2015-03-26 |
| | Asian | 1418 | Human | Morelos | 2015-04-14 |
| | Asian | 4124 | Human | Chiapas | 2015-08-12 |
| | Asian | 2135 | Human | Veracruz | 2015-06-08 |
| | Asian | 1825 | Human | Veracruz | 2015-05-16 |
| | Asian | 3430 | Human | Quintana Roo | 2015-07-23 |
| | Asian | 2007 | Human | Michoacan | 2015-05-13 |
| | Asian | 2430 | Human | Michoacan | 2015-06-05 |
| | Asian | 5439 | Human | Guerrero | 2015-11-20 |
| | Asian | 1343 | Human | Guerrero | 2015-04-10 |

#### Table S2. PAHO regions

| PAHO region | Country |
|---|---|
| North America | USA |
| North America | Mexico |
| North America | Canada |
| North America | Bermuda |
| Central America | Panama |
| Central America | Nicaragua |
| Central America | Honduras |
| Central America | Guatemala |
| Central America | El Salvador |
| Central America | Costa Rica |
| Central America | Belize |
| Latin America | Saint Martin |
| Latin America | Martinique |
| Latin America | Guadeloupe |
| Latin America | Puerto Rico |
| Latin America | Haiti |
| Latin America | Guyana |
| Latin America | French Guyana |
| Latin America | Dominican Republic |
| Latin America | Cuba |
| Andean | Venezuela |
| Andean | Peru |
| Andean | Ecuador |
| Andean | Colombia |
| Andean | Bolivia |
| South Cone | Uruguay |
| South Cone | Paraguay |
| South Cone | Chile |
| South Cone | Brazil |
| South Cone | Argentina |
| Non-Latin Caribbean | Saint Barthelemy |
| Non-Latin Caribbean | Sint Maarten |
| Non-Latin Caribbean | United States Virgin Islands |
| Non-Latin Caribbean | Turks and Caicos Islands |
| Non-Latin Caribbean | Trinidad and Tobago |
| Non-Latin Caribbean | Suriname |
| Non-Latin Caribbean | Saint Vincent and the Grenadines |
| Non-Latin Caribbean | Saint Lucia |
| Non-Latin Caribbean | Saint Kitts and Nevis |
| Non-Latin Caribbean | Montserrat |
| Non-Latin Caribbean | Jamaica |
| Non-Latin Caribbean | Grenada |
| Non-Latin Caribbean | Dominica |
| Non-Latin Caribbean | Curacao |
| Non-Latin Caribbean | Cayman Islands |
| Non-Latin Caribbean | British Virgin Islands |
| Non-Latin Caribbean | Bonaire Saint Eustatius and Saba |
| Non-Latin Caribbean | Barbados |
| Non-Latin Caribbean | Bahamas |
| Non-Latin Caribbean | Aruba |
| Non-Latin Caribbean | Antigua and Barbuda |
| Non-Latin Caribbean | Anguilla |

#### Table S3. BSSVS results for CHIKV

| FROM | TO | BAYES_FACTOR | POSTERIOR PROBABILITY |
|---|---|---|---|
| Americas | Centre-north | 86.20856163 | 0.922105978 |
| Americas | Centre-south | 0.191914894 | 0.025676635 |
| Americas | East | 0.743297742 | 0.092614881 |
| Americas | Northeast | 0.868885317 | 0.106595068 |
| Americas | Northwest | 0.177265425 | 0.023763232 |
| Americas | Southeast | 1.062599441 | 0.127333889 |
| Americas | Southwest | 5236.520985 | 0.99861124 |
| Americas | West | 15.76167184 | 0.683979878 |
| Centre-north | Centre-south | 1.060829123 | 0.127148721 |
| Centre-north | East | 1.07975133 | 0.129123847 |
| Centre-north | Northeast | 1.438552475 | 0.164953862 |
| Centre-north | Northwest | 1.671856523 | 0.186711107 |
| Centre-north | Southeast | 0.989757074 | 0.119649415 |
| Centre-north | Southwest | 1.039937246 | 0.124957566 |
| Centre-north | West | 1.07708521 | 0.128846094 |
| Centre-south | East | 0.816846997 | 0.100854859 |
| Centre-south | Northeast | 8.62935726 | 0.542326328 |
| Centre-south | Northwest | 12.1567072 | 0.625374194 |
| Centre-south | Southeast | 0.796051295 | 0.098540259 |
| Centre-south | Southwest | 0.725594228 | 0.090608894 |
| Centre-south | West | 0.743024787 | 0.09258402 |
| East | Northeast | 2.684716558 | 0.269357776 |
| East | Northwest | 7.099083565 | 0.493627133 |
| East | Southeast | 1.128933359 | 0.134215968 |
| East | Southwest | 0.940751488 | 0.114402987 |
| East | West | 0.880728587 | 0.107891245 |
| Northeast | Northwest | 4.998184013 | 0.406999352 |
| Northeast | Southeast | 2.498801692 | 0.255470173 |
| Northeast | Southwest | 1.712473846 | 0.190383606 |
| Northeast | West | 0.956254924 | 0.1160695 |
| Northwest | Southeast | 1.257392275 | 0.147239453 |
| Northwest | Southwest | 1.40068558 | 0.161312224 |
| Northwest | West | 1.255847284 | 0.147085146 |
| Southeast | Southwest | 2.554663903 | 0.259698176 |
| Southeast | West | 1.126535435 | 0.133969077 |
| Southwest | West | 193.5441261 | 0.963737926 |
| Centre-north | Americas | 1.603408292 | 0.180446255 |
| Centre-south | Americas | 21.97990965 | 0.751134154 |
| East | Americas | 1.081826155 | 0.129339876 |
| Northeast | Americas | 1.553499315 | 0.175817054 |
| Northwest | Americas | 2.00818265 | 0.216152825 |
| Southeast | Americas | 1.362512639 | 0.157608863 |
| Southwest | Americas | 1.233353924 | 0.144832269 |
| West | Americas | 0.939891896 | 0.114310403 |
| Centre-south | Centre-north | 17.4214616 | 0.70521248 |
| East | Centre-north | 0.880728587 | 0.107891245 |
| Northeast | Centre-north | 1.367900054 | 0.158133506 |
| Northwest | Centre-north | 1.733093224 | 0.192235287 |
| Southeast | Centre-north | 1.121743685 | 0.133475295 |
| Southwest | Centre-north | 1.464737596 | 0.167453631 |
| West | Centre-north | 0.795221687 | 0.098447675 |
| East | Centre-south | 0.947634694 | 0.11514366 |
| Northeast | Centre-south | 1.27318317 | 0.148813381 |
| Northwest | Centre-south | 1.448232368 | 0.165879702 |
| Southeast | Centre-south | 1.248439107 | 0.146344474 |
| Southwest | Centre-south | 10.16593938 | 0.582631238 |
| West | Centre-south | 26.55345456 | 0.784773015 |
| Northeast | East | 3.538504216 | 0.327006759 |
| Northwest | East | 1.238581511 | 0.145356911 |
| Southeast | East | 4.83050106 | 0.398790235 |
| Southwest | East | 7.167776439 | 0.496034318 |
| West | East | 38.72485751 | 0.841712187 |
| Northwest | Northeast | 1.565426269 | 0.176928062 |
| Southeast | Northeast | 5.627732872 | 0.435916427 |
| Southwest | Northeast | 15.5321893 | 0.68080116 |
| West | Northeast | 0.824916734 | 0.101749838 |
| Southeast | Northwest | 1.907953351 | 0.207604234 |
| Southwest | Northwest | 2.099045763 | 0.223744715 |
| West | Northwest | 0.754778625 | 0.093911058 |
| Southwest | Southeast | 32.14470707 | 0.81529488 |
| West | Southeast | 2.949647346 | 0.288275777 |
| West | Southwest | 0.916750669 | 0.111810635 |

#### Table S4. BSSVS results for DENV-2

| FROM | TO | BAYES_FACTOR | POSTERIOR PROBABILITY |
|---|---|---|---|
| Belize | Centre-south | 0.967626508 | 0.133524069 |
| Belize | East | 0.953020166 | 0.131774117 |
| Belize | Guatemala | 0.957650717 | 0.132329658 |
| Belize | Nicaragua | 1.70136634 | 0.213188523 |
| Belize | Southeast | 0.930185929 | 0.129024194 |
| Belize | Southwest | 1.025702329 | 0.140412766 |
| Belize | West | 0.942161662 | 0.130468598 |
| Centre-south | East | 1.124996057 | 0.151940224 |
| Centre-south | Guatemala | 1.150059576 | 0.154801256 |
| Centre-south | Nicaragua | 1.410877431 | 0.183467126 |
| Centre-south | Southeast | 1.105886879 | 0.14974584 |
| Centre-south | Southwest | 1.234011655 | 0.164245438 |
| Centre-south | West | 1.001469487 | 0.137551735 |
| East | Guatemala | 1.130092361 | 0.152523541 |
| East | Nicaragua | 1.293408656 | 0.170800811 |
| East | Southeast | 1.210612198 | 0.161634399 |
| East | Southwest | 1.173818724 | 0.157495625 |
| East | West | 1.064144217 | 0.144912641 |
| Guatemala | Nicaragua | 1.391307134 | 0.18138385 |
| Guatemala | Southeast | 1.126936679 | 0.15216244 |
| Guatemala | Southwest | 1.159593991 | 0.155884559 |
| Guatemala | West | 1.146886867 | 0.154440154 |
| Nicaragua | Southeast | 0.338795477 | 0.051193022 |
| Nicaragua | Southwest | 1124.009198 | 0.994444599 |
| Nicaragua | West | 0.263140384 | 0.040221105 |
| Southeast | Southwest | 1.257557779 | 0.166856476 |
| Southeast | West | 1.201194161 | 0.160578873 |
| Southwest | West | 2929.534829 | 0.997861171 |
| Centre-south | Belize | 1.714064821 | 0.214438488 |
| East | Belize | 2.004936908 | 0.242021055 |
| Guatemala | Belize | 2.192768344 | 0.258826144 |
| Nicaragua | Belize | 4.066210204 | 0.393044638 |
| Southeast | Belize | 2.42268372 | 0.278408933 |
| Southwest | Belize | 3.166627625 | 0.335240688 |
| West | Belize | 1.245766298 | 0.165550957 |
| East | Centre-south | 1.082558193 | 0.147051471 |
| Guatemala | Centre-south | 1.105404382 | 0.149690286 |
| Nicaragua | Centre-south | 0.251233526 | 0.038471154 |
| Southeast | Centre-south | 1.132521639 | 0.152801311 |
| Southwest | Centre-south | 0.754762935 | 0.107302575 |
| West | Centre-south | 83.24858871 | 0.929863059 |
| Guatemala | East | 1.233262606 | 0.164162107 |
| Nicaragua | East | 0.27129225 | 0.041415516 |
| Southeast | East | 2.168390841 | 0.256687314 |
| Southwest | East | 56.08153389 | 0.899308353 |
| West | East | 0.749951141 | 0.106691481 |
| Nicaragua | Guatemala | 0.823495905 | 0.115941224 |
| Southeast | Guatemala | 1.282010601 | 0.169550846 |
| Southwest | Guatemala | 134.6545105 | 0.955445682 |
| West | Guatemala | 0.753668767 | 0.10716369 |
| Southeast | Nicaragua | 1.480033803 | 0.190744702 |
| Southwest | Nicaragua | 1.196246807 | 0.160023333 |
| West | Nicaragua | 0.800140535 | 0.113024638 |
| Southwest | Southeast | 1270.882832 | 0.99508347 |
| West | Southeast | 0.713638796 | 0.102052721 |
| West | Southwest | 0.753231195 | 0.107108136 |
